# U.S. Field Hospitals: A Study on Public Health Emergency Response to COVID-19

**DOI:** 10.1101/2020.08.05.20169094

**Authors:** Luorongxin Yuan, Sherryn Sherryn, Peter Hu, Fenghao Chen

## Abstract

With the number of confirmed COVID-19 cases rapidly growing in the U.S., many states are experiencing a shortage of hospital—especially ICU—beds. In addition to discharging non-critical patients, expanding local hospitals’ capacity as well as re-opening closed healthcare facilities, these states are actively building or converting public venues into field hospitals to fill the gap^1^. By studying these makeshift hospitals, we found that the states most severely impacted by the pandemic are fast at responding with the first wave of hospitals opening around the date of peak demand and the majority ready to use by the end of April. However, depending on the types of patients the field hospitals accept (COVID-19 vs. non-COVID-19) and how they are incorporated to local healthcare system, these field hospitals have utilization rate ranging from 100% to 0%. The field hospitals acting as alternative site to treat non-COVID-19 patients typically had low utilization rate and often faced the risk of COVID-19 outbreak in the facility. As overflow facilities, the field hospitals providing intensive care were highly relied on by local healthcare systems whereas the field hospitals dedicated to patients with mild symptoms often found it hard to fill the beds due to a combination of factors such as strict regulation on transferring patients from local hospitals, complication of health insurance discouraging health-seeking behavior, and effective public health measure to “flatten the curve” so that the additional beds were no longer needed.

## Introduction

The first COVID-19 in the U.S. was confirmed on Jan. 21^st^, but local transmission was not reported until Feb. 26^th^. From early March, the spreading of COVID-19 began to accelerate and by Mar. 13^th^, President Donald Trump declared National Emergency, shortly after which COVID-19 cases were reported in all 50 states, District of Columbia and a few territories^2^. To monitor the global spreading of COVID-19 live, the Center for System Sciences and Engineering at the Johns Hopkins University (JHU CSSE) launched the COVID tracking map on Jan 21^st^ and when the number of accumulative cases became an alarming 467.8K in the U.S., the Institute for Health Metrics and Evaluation at the University of Washington (IHME) released a projection model on Mar. 25^th^ to predict the surge of affected population and accordingly the imminent medical crisis^3-4^. Seeing the rapid growth of confirmed cases in each state and the potential medical supply shortage, almost all state government took the initiative to recruit available beds and seek public venues to build makeshift hospitals. Even though there is mass media coverage on the building and opening of these field hospitals, little is known about how the decision was made and how wise they are—namely, whether the field hospitals are effectively sharing the load with local hospitals. In this study, we hope to answer 1) if the field hospitals opened in time ready for the surge, 2) if their capacity met the predicted demand, 3) what particular role they play in the medical system, and 4) if they have fulfilled their intended purpose to relieve the stress experienced by local hospitals.

## Method

Information regarding hospital capacities in the U.S. was obtained from American Hospital Directory, Inc. online database through JHU subscription^5^. All data points about field hospitals were collected manually by searching keywords such as “COVID-19”, “coronavirus”, “field hospital” online. The information collected from the news feeds include the location of field hospitals, construction time, opening date, type of patients they accept (COVID-19 vs. non-COVID-19), affiliation (military vs. non-military) and updates on how they are running. Unless noted otherwise, the operational date is estimated as one day after the construction end date. Information about Army-built field hospital was obtained from Army Corps of Engineers website^6^. Live update on confirmed cases, incidence rate, hospitalization rate, etc. was downloaded from the JHU CSSE COVID GitHub and the COVID-19 estimate dataset was obtained from IHME COVID-19 projection open results^3-4^. The update cutoff date is May 1^st^.

## Results

Two weeks into National Emergency, two United States Naval Ship *(USNS)* medical cruise first docked in Los Angeles and New York City, the two epicenters at the time, followed by the construction of more than 70 field hospitals in 24 states, supplying more than 27K beds in the next 2 months **(Table 1)**. About half of the field hospitals started operating while the rest remained closed until further notice. The majority of the field hospitals take COVID-19 patients with mild symptoms and were built by Army Corps of Engineers, National Guards, and volunteers. The first field batch of field hospitals started receiving patients on April 1^st^; taking into consideration that it takes 1-2 weeks on average to prepare one field hospital, many states have taken actions immediately after the declaration of National Emergency^1^.

**Table 1:** List of Existing and Planned Field Hospital & Shelters^21-69^. All dates are in 2020.

| Hospital Name | Beds | COVID | Military | City | State | Peak Demand | Constr. Start | Constr. End | Operational Date |
| --- | --- | --- | --- | --- | --- | --- | --- | --- | --- |
| Oregon Convention Center Shelter | 140 | No | No | Portland | OR | 4/9 |  |  | 3/21 |
| USNS Mercy | 1000 | No | Yes | San Pedro | CA | 4/17 |  |  | 3/27 |
| USNS Comfort | 1000 | No | Yes | NYC | NY | 4/8 |  |  | 3/30 |
| San Diego Convention Center Shelter | 400 | No | No | San Diego | CA | 4/17 |  |  | 4/1 |
| Western Connecticut State University O'Neill Center | 200 | Yes | No | Danbury | CT | 4/10 |  |  | 4/1 |
| Central Park Field Hospital | 68 | Yes | No | NYC | NY | 4/8 |  |  | 4/1 |
| Shorline Soccer Field | 200 | Yes | No | Shorline | WA | 4/19 |  |  | 4/2 |
| Javits New York Medical Station | 1000 | Yes | Yes | NYC | NY | 4/8 | 3/30 | 4/8 | 4/4 |
| Ernest N. Morial Convention Center Medical Center | 1000 | Yes | No | New Orleans | LA | 4/14 |  |  | 4/6 |
| CenturyLink | 250 | No | Yes | Seattle | WA | 4/19 |  |  | 4/6 |
| Colorado Convention Center Shelter | 600 | No | Yes | Denver | CO | 4/28 | 4/9 | 4/24 | 4/7 |
| Saint Francis Field Hospital | 25 | Yes | Yes | Hartford | CT | 4/10 |  |  | 4/7 |
| Santa Clara Convention Center | 250 | Yes | Yes | Santa Clara | CA | 4/17 |  |  | 4/8 |
| Danbury Field Hospital | 25 | Yes | Yes | Danbury | CT | 4/10 |  |  | 4/8 |
| Middlesex Health Field Hospital | 25 | Yes | Yes | Middletown | CT | 4/10 |  |  | 4/8 |
| DCU Center | 250 | Yes | No | Worcester | MA | 4/12 |  |  | 4/9 |
| Southern Connecticut State University Moore Fieldhouse | 250 | Yes | Yes | New Haven | CT | 4/10 |  |  | 4/10 |
| McCormick Place | 1000 | Yes | Yes | Chicago | IL | 4/8 | 3/29 | 4/24 | 4/10 |
| Boston Convention and Exposition Center | 1000 | Yes | No | Boston | MA | 4/12 |  |  | 4/10 |
| Newtown Athletic Club | 300 | No | No | Newtown | PA | 4/17 |  |  | 4/10 |
| TCF Center Field Hospital | 1000 | Yes | Yes | Detroit | MI | 4/8 | 4/1 | 4/9 | 4/11 |
| New Jersey Convention and Exposition Center | 500 | Yes | Yes | Edison | NJ | 4/9 |  |  | 4/11 |
| Sharon Field Hospital | 25 | Yes | Yes | Sharon | CT | 4/10 |  |  | 4/12 |
| Connecticut Convention Center | 646 | Yes | Yes | Hartford | CT | 4/10 |  |  | 4/15 |
| Missouri ACF Quality Inn at Florissant | 120 | Yes | Yes | Florissant | MO | 4/28 | 4/8 | 4/11 | 4/15 |
| Meadowlands Exposition Center | 250 | Yes | No | Secaucus | NJ | 4/9 |  |  | 4/15 |
| Craneway Pavilion | 250 | Yes | Yes | Richmond | CA | 4/17 |  |  | 4/17 |
| Alaska Airlines Center | 163 | Yes | Yes | Anchorage | AK | 4/18 | 4/9 | 4/17 | 4/18 |
| Westchester Community Center | 100 | Yes | Yes | White Plains | NY | 4/8 | 3/27 | 4/17 | 4/18 |
| Wisconsin State Fair | 530 | Yes | Yes | West Allis | WI | 4/14 | 4/8 | 4/18 | 4/19 |
| Miami Beach Convention Center | 450 | Yes | Yes | Miami Beach | FL | 5/3 | 4/8 | 4/19 | 4/20 |
| Atlantic City Convention Center | 250 | No | Yes | Atlantic City | NJ | 4/9 |  |  | 4/20 |
| Miyamura High School | 50 | Yes | Yes | Gallup | NM | 4/25 | 4/6 | 4/19 | 4/20 |
| New Bridge-Bergen Medical Center | 40 | No | Yes | Paramus | NJ | 4/9 | 4/9 | 4/22 | 4/23 |
| Suburban Collection Showplace | 250 | Yes | Yes | Novi | MI | 4/8 | 4/6 | 4/20 | 4/24 |
| East Orange Hospital | 250 | Yes | Yes | East Orange | NJ | 4/9 | 4/9 | 4/23 | 4/24 |
| New Bridge Hospital | 40 | Yes | Yes | Paramus | NJ | 4/9 | 4/9 | 4/23 | 4/24 |
| Metro South Medical Ctr | 350 | Yes | Yes | Blue Island | IL | 4/8 | 3/30 | 4/24 | 4/25 |
| Sherman Hospital | 283 | Yes | Yes | Eglin | IL | 4/8 | 3/30 | 4/24 | 4/25 |
| Westlake Hospital | 314 | Yes | Yes | Melrose Park | IL | 4/8 | 4/5 | 4/24 | 4/25 |
| Hagerstown Correctional Facility | 192 | Yes | Yes | Hagerstown | MD | 4/11 | 4/13 | 4/24 | 4/25 |
| Baltimore Convention Center | 250 | Yes | Yes | Baltimore | MD | 4/11 |  | 4/13 | 4/27 |
| SUNY Old Westbury | 1022 | Yes | Yes | Old Westbury | NY | 4/8 | 3/31 | 4/26 | 4/27 |
| SUNY Stony Brook | 1038 | Yes | Yes | Stony Brook | NY | 4/8 | 3/29 | 4/26 | 4/27 |
| St. Francis Hospital | 37 | Yes | Yes | Trenton | NJ | 4/9 | 4/14 | 4/27 | 4/28 |
| The Ranch Events Complex | 1007 | Yes | Yes | Loveland | CO | 4/28 | 4/9 | 4/29 | 4/30 |
| New Bridge-Bergen Med Ctr Parking Lot | 100 | Yes | Yes | Paramus | NJ | 4/9 | 4/15 | 4/29 | 4/30 |
| Chinle Community Center | 50 | No | Yes | Chinle | AZ | 4/30 | 4/18 | 5/1 | 5/2 |
| Atsa Biyaazh Community Center | 40 | No | Yes | Shiprock | NM | 4/25 | 4/18 | 5/1 | 5/2 |
| Eugene River Facility | 42 | Yes | Yes | Eugene | OR | 4/9 | 4/18 | 5/1 | 5/2 |
| Walter Washington Community | 491 | Yes | Yes | Washington | DC | 4/26 | 4/18 | 5/8 | 5/9 |
| St. Luke's Medical Center | 411 | Yes | Yes | Phoenix | AZ | 4/30 | 4/16 | 5/14 | 5/15 |
| United Medical Center (UMC) | 6 | Yes | Yes | Washington | DC | 4/9 | 4/9 | 4/22 | 5/15 |
| Commercial Appeal Building | 170 | Yes | Yes | Memphis | TN | 4/16 | 4/9 | 5/14 | 5/15 |
| Dulles Expo Center | 510 | TBD | Yes | Chantilly | VA | 5/2 |  |  | 5/15 |
| Van Cortland Park | 200 | Yes | No | The Bronx | NY | 4/8 |  |  | canceled |
| Kay Bailey Hutchison Convention Center Medical Center | 1400 | Yes | Yes | Dallas | TX | 4/18 |  |  | canceled |
| Hampdon Roads Convention Center | 580 | TBD |  | Hampton | VA | 5/2 |  |  | canceled |
| Los Angeles Convention Center | 250 | Yes | Yes | Los Angeles | CA | 4/17 |  |  | NA |
| Miami-Dade County Fairground | 250 | Yes | No | Miami | FL | 5/3 |  |  | NA |
| South Florida Fairgrounds | 200 | Yes | No | West Palm Beach | FL | 5/3 | 4/7 |  | NA |
| Dalton Convention Center | 150 |  | No | Dalton | GA | 4/21 |  |  | NA |
| Joint Base Cape Cod | 150 | Yes | Yes | Bourne | MA | 4/12 |  |  | NA |
| Dayton Convention Center |  | Yes | No | Dayton | OH | 4/16 |  |  | NA |
| Sea Gate Convention Center | 415 | Yes | No | Toledo | OH | 4/16 |  |  | NA |
| East Stroudsburg Campus |  |  | No | Stroudsburg | PA | 4/17 |  |  | NA |
| Music City Center |  | Yes | Yes | Nashville | TN | 4/16 |  |  | NA |
| Richmond Convention Center | 758 | TBD | Yes | Richmond | VA | 5/2 |  |  | NA |
| Porterville Dev. Center | 246 | Yes | Yes | Porterville | CA | 4/17 | 4/8 | 4/21 | TBD |
| San Mateo Event Center | 250 | No | No | San Mateo | CA | 4/17 |  |  | TBD |
| Gibson Medical Center | 200 | Yes | Yes | Albuquerque | NM | 4/25 | 4/3 | 4/17 | TBD |
| Case Western Health Education Campus Hope Hospital | 327 | Yes | No | Cleveland | OH | 4/16 |  | 4/2 | TBD |
| Covelli Convention Center | 200 | Yes | Yes | Youngstown | OH | 4/16 |  |  | TBD |
| Duke Energy Convention Center | 500 | Yes | No | Cincinnati | OH | 4/16 |  | 4/16 | TBD |
| Greater Columbus Convention Center | 1000 | Yes | No | Columbus | OH | 4/16 |  | 4/14 | TBD |

### Field hospitals are concentrated in states with International airports and high population density

First, we mapped the existing and planned field hospitals to visualize their spatial distribution **(Figure 1)**. It was not surprising to see that the field hospitals clustered around major international airports, such as San Francisco, Los Angeles, Seattle, Chicago, Dallas, and New York, that have connections to China—where the first epicenter was—and to Europe—where most transatlantic import occurred—especially before the international travel restriction was published on the 31^st^ of March^7-8^. In addition to imported cases, the coronavirus then spread out to other cities through domestic travels and community-based transmission. Where the field hospitals indexed the most affected states, their distribution highly resembles that of imported measles outbreak predicted according to 1) international travel volume and port of entry, 2) incidence rate at the origin, and 3) the population of surrounding county^9^. If an international epidemic originating from Europe or Asia occurs in the future, these states should the first ones to start public health emergency response procedures.

**Figure 1:**
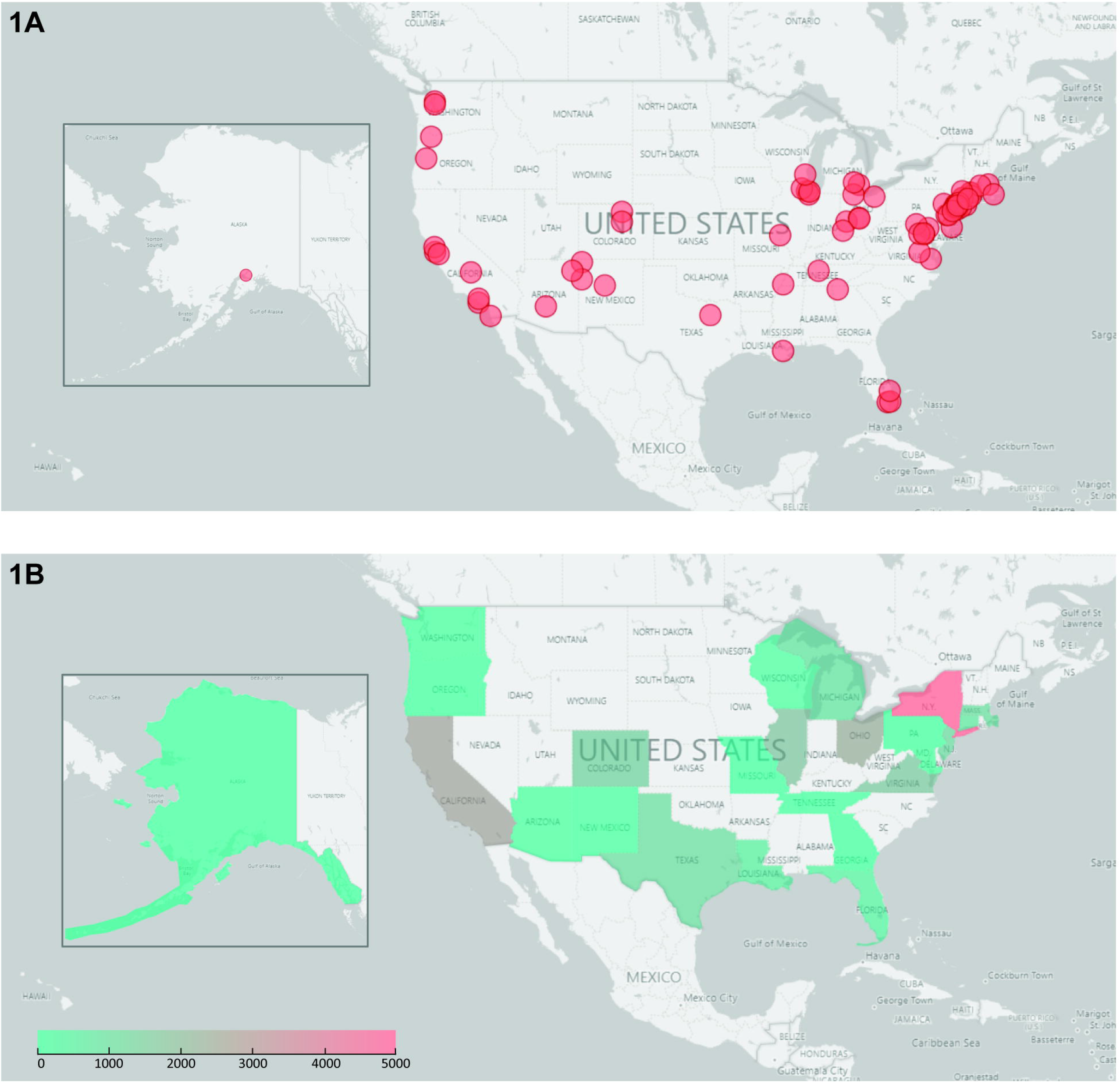
Visualizing the spatial distribution of field hospitals. **1A:** Current and planned field hospitals and shelters mapped to their location using Power BI. Each dot represents one field hospital regardless of its capacity. **1B:** Density map indicating total number of field hospital beds in each state.

Next, the question becomes, when the local epidemics developed into national pandemic, what determined when and where to build field hospitals? While reading through news reports, it was hard to ignore that the building and opening of field hospitals is a very dynamic process to accommodate the evolving curve of existing confirmed cases. Besides the field hospitals affiliated to and staffed by local healthcare system, recruiting staff can be challenging during the pandemic. Therefore, many field hospitals were constructed but remained closed until necessary to reserve medical resources. Similarly, in states where the outbreak was not as urgent, the potential sites were outfitted but no construction would start until situation worsens; alternatively, if the situation improves the construction would be canceled. Therefore, the decision must be dependent on the live feeds of the growing numbers. For convenience, we collected the U.S. COVID-19 statistics on April 15th, the national peak predicted by IHME, shown in **Table 2**.

**Table 2:** Logistics and JHU CSSE COVID-19 updates of states that have existing or planned field hospitals. All COVID-19 data below reflect the outbreak on April 15^th^, the average peak date nationwide. Incidence rate is the number of confirmed cases per 100,000 persons. The ranking number is assigned to each state when either accumulative confirmed cases or incidence rate of all 50 states and Washington DC are ranked from largest to smallest. Land area is measured in mi^2^, and population density in persons/mi^2^. Available bed counts reflect hospital capacity in each state before the COVID-19 outbreak. Bed rate is calculated as number of beds available per 100,000 persons.

| State | Confirmed | Ranking | Deaths | Active | Incidence Rate | Ranking | Field Hospital Bed | Population | Land Area | Population Density | Available Bed | Bed Rate |
| --- | --- | --- | --- | --- | --- | --- | --- | --- | --- | --- | --- | --- |
| Alaska | 293 | 50 | 9 | 284 | 49.0 | 47 | 163 | 731545 | 570641 | 1.3 | 683 | 93.4 |
| Arizona | 3964 | 23 | 142 | 3822 | 54.5 | 44 | 461 | 7278717 | 113594 | 64.1 | 6018 | 82.7 |
| California | 26686 | 6 | 860 | 25826 | 68.1 | 35 | 2646 | 39512223 | 155779 | 253.6 | 26654 | 67.5 |
| Colorado | 7956 | 16 | 328 | 7628 | 140.4 | 16 | 1607 | 5758736 | 103642 | 55.6 | 4852 | 84.3 |
| Connecticut | 14755 | 12 | 868 | 13887 | 413.9 | 5 | 1196 | 3565278 | 4842 | 736.3 | 1818 | 51.0 |
| Florida | 22511 | 8 | 596 | 21915 | 106.0 | 21 | 900 | 21477737 | 53625 | 400.5 | 20184 | 94.0 |
| Georgia | 14987 | 11 | 552 | 14435 | 147.8 | 14 | 150 | 10617423 | 57513 | 184.6 | 8323 | 78.4 |
| Illinois | 24593 | 7 | 949 | 23644 | 209.6 | 10 | 1947 | 12671821 | 55519 | 228.2 | 14552 | 114.8 |
| Indiana | 8960 | 15 | 436 | 8524 | 136.9 | 17 | 0 | 6732219 | 35826 | 187.9 | 8485 | 126.0 |
| Louisiana | 21951 | 9 | 1103 | 20848 | 477.5 | 3 | 1000 | 4648794 | 43204 | 107.6 | 7205 | 155.0 |
| Maryland | 10032 | 14 | 311 | 9721 | 168.8 | 12 | 442 | 6045680 | 9707 | 622.8 | 3961 | 65.5 |
| Massachusetts | 29918 | 3 | 1108 | 28810 | 435.9 | 4 | 1400 | 6892503 | 7800 | 883.7 | 4849 | 70.4 |
| Michigan | 28059 | 4 | 1921 | 26138 | 352.2 | 6 | 1250 | 9986857 | 56539 | 176.6 | 10155 | 101.7 |
| Missouri | 4791 | 21 | 153 | 4638 | 81.8 | 29 | 120 | 6137428 | 68742 | 89.3 | 7933 | 129.3 |
| New Jersey | 71030 | 2 | 3156 | 67874 | 799.7 | 2 | 1467 | 8882190 | 7354 | 1207.8 | 7815 | 88.0 |
| New Mexico | 1484 | 39 | 36 | 1448 | 89.0 | 23 | 290 | 2096829 | 121298 | 17.3 | 1753 | 83.6 |
| New York | 214454 | 1 | 11617 | 202837 | 1271.9 | 1 | 4428 | 19453561 | 47126 | 412.8 | 13011 | 66.9 |
| Ohio | 7794 | 17 | 362 | 7432 | 69.8 | 33 | 2442 | 11689100 | 40861 | 286.1 | 14291 | 122.3 |
| Oregon | 1663 | 36 | 58 | 1605 | 41.5 | 49 | 182 | 4217737 | 95988 | 43.9 | 2658 | 63.0 |
| Pennsylvania | 26753 | 5 | 779 | 25974 | 212.5 | 9 | 300 | 12801989 | 44743 | 286.1 | 14395 | 112.4 |
| Tennessee | 5827 | 19 | 124 | 5703 | 88.8 | 25 | 170 | 6829174 | 41235 | 165.6 | 7812 | 114.4 |
| Texas | 15907 | 10 | 375 | 15532 | 69.2 | 34 | 1400 | 28995881 | 261232 | 111.0 | 28634 | 98.8 |
| Virginia | 6500 | 18 | 195 | 6305 | 82.2 | 28 | 1848 | 8535519 | 39490 | 216.1 | 6581 | 77.1 |
| Washington | 10942 | 13 | 552 | 10390 | 144.9 | 15 | 450 | 7614893 | 66456 | 114.6 | 4907 | 64.4 |
| Washington DC | 2197 | 32 | 72 | 2125 | 311.3 | 7 | 497 | 702455 | 68.34 | 10278.8 | 1094 | 155.7 |
| Wisconsin | 3721 | 24 | 183 | 3538 | 71.9 | 32 | 530 | 5822434 | 97093 | 60.0 | 5365 | 92.1 |

### The field hospital opening is more Influenced by accumulated confirmed cases in each state

The timeline histogram shows that the greatest number of field hospitals opened during the 5^th^ week, where most state met bed demand peaks **(Table 1, Fig 2A)**. Intuitively, the more urgent the COVID-19 outbreak was in the state, the earlier the field hospitals would be built. For COVID-19, the recovery time is calculated from the first day showing symptom or having tested positive to the last day of testing negative for two consecutive days^10-11^. During the early phase of highly contagious disease with doubling time of 5 days and recovery time of 10 days, the recovery and death rate is almost negligible, and it is essential to contract trace, identify and isolate the infected individuals to slow down the spreading^12^. Therefore, the confirmed cases and the incidence rate, defined as number of infected individuals per 100,000 persons, should be more correlated with the opening time of field hospitals. Because larger population will produce larger number of confirmed cases, it is perceived that the population-independent infected rate would be a more accurate indicator of when to build the field hospitals. However, when we indexed each state with a ranking number, where either accumulative confirmed case number or incidence rate of all 50 state and Washington DC on Apr. 15^th^ was ranked from largest to smallest, and plotted against the opening date of the field hospitals, a stronger correlation was observed between ranking of accumulative confirmed cases and the field hospital opening date **(Fig 2B and D)**. For example, California had an alarming number of accumulative confirmed cases of 26,686, ranking 6^th^ in the nation; however, by incidence rate it was ranked 35^th^, indicating that the outbreak in the state was not as bad as it appeared even though the incidence rate was exceptionally high in certain cities. At the state level, the additional field hospital beds seemed unnecessary but at city level, they were essential. Consequently, a more case-specific micro-scale analysis is required to describe the discrepancy between the ranking of accumulative confirmed cases and of incidence rate. This observation could also be contributed to the fact that JHU CSSE did not publish the county-level population-corrected COVID-19 outbreak until Apr. 12^th^. Even though the incidence rate could be calculated with simple math and more reliable, the mounting number of accumulative confirmed cases may have had a heavier impact on decision-making psychologically.

**Figure 2:**
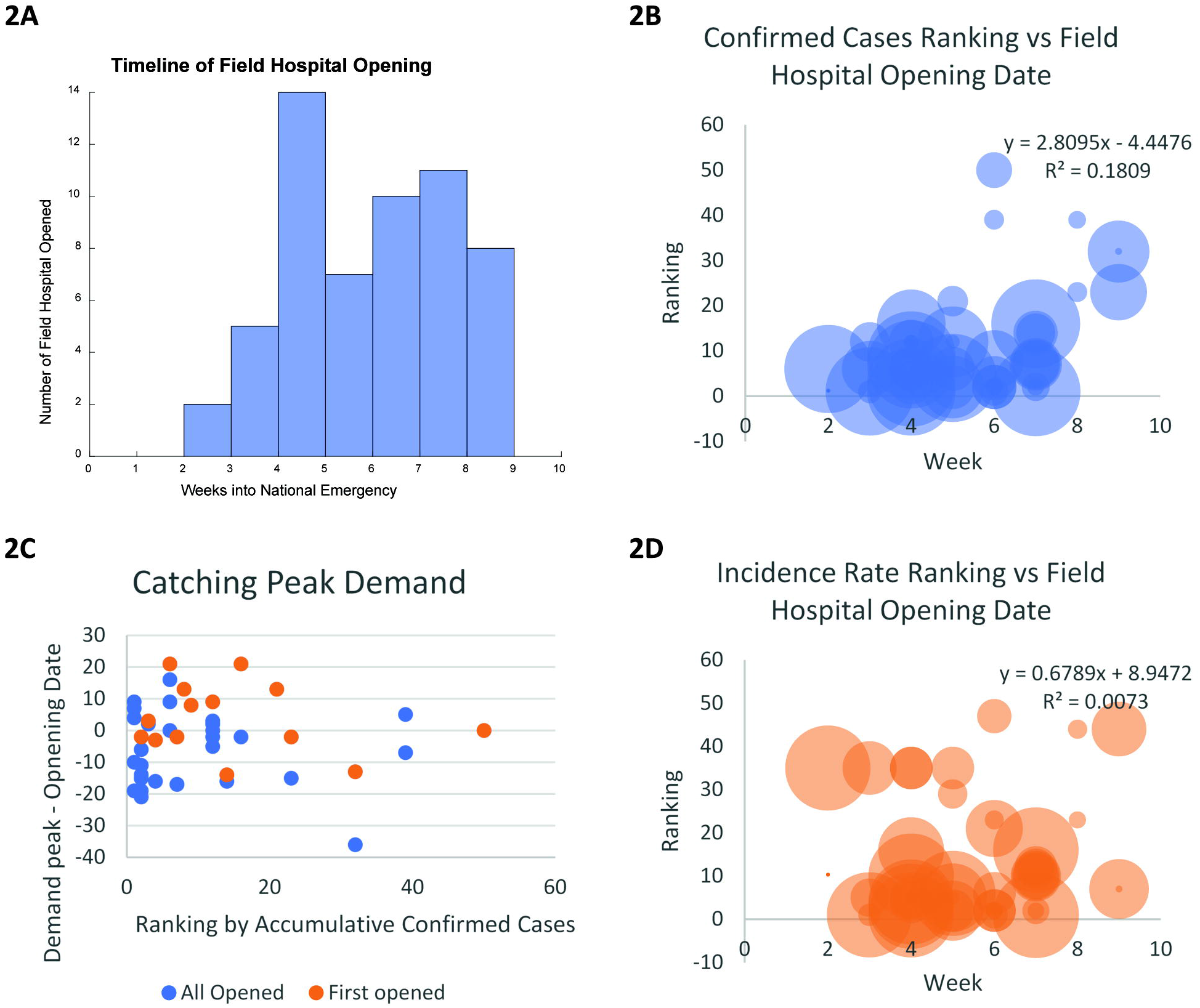
The accumulative confirmed cases is a more important factor determining when and where to build the field hospitals. **2A:** Histogram of field hospital opening time, binned to weeks into National Emergency. **2B and D:** Urgency level, approximated by accumulated confirmed cases or incidence rate ranking, against opening date of field hospitals with linear fit. Radii of circles indicate the capacity of each field hospital. **2C:** Most opening dates of field hospitals are clustered around y=0 or the date of predicted peak demand in the state they belong to. The first field hospital in each state on average opened before or by peak demand date. Each dot represents one field hospital. All field hospitals shown in blue, the first field hospital to open in each state shown in orange.

### Field hospitals in each state opened promptly to meet peak demand

Then we ask the question—were the field hospitals built by the date of peak demand? Knowing that the accumulative confirmed number is the determining factor of building these field hospitals, we plotted the difference between peak demand peak and field hospital opening date against the ranking of accumulative confirmed cases **(Fig 2C)**. A positive y-value indicates that the field hospital opened before demand peak arrived, whereas a negative value suggests otherwise. From the scatter plot, we could conclude that most hospitals were opened around the day of peak demand. When the first hospital in each state to open was highlighted in orange, we could see that on average they were put to service well before or around the day of peak demand. It is noteworthy that some larger field hospitals, such as the Javits New York Medical Station, opened before construction ended to account for the imminent bed shortage **(Table 1)**. In general, field hospitals in each state opened promptly to meet peak demand.

### The total number of beds in field hospitals is most correlated to the difference between incidence rate and bed rate in each state

In parallel to the question when and where to build the field hospital is how many additional beds are needed? The classic SIR (Susceptible-Infected-Recovered) model of infectious disease suggests that the maximal infected rate *I_max_/N* can be estimated as

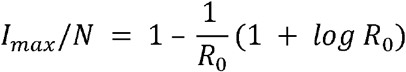

where *I_max_* is the maximal number of infected individuals, *N* the population in the defined area, and *R*_0_ the basic reproduction number of the infectious disease^13^. At first *R*_0_ was estimated to be 3.65, which means that on average, 2.55 individuals will be infected by any given infected person, however after the implementation of social distancing, the effective *R*_0_ is estimated to be 2.55 for COVID-19 in the U.S.^12^. Because the number of available hospital beds is directly proportional to population **(Fig 3A)**, and the hospitalization rate (number of patients hospitalized over number of confirmed cases) is known to each state, the maximal number of additional beds required should be more or less proportional to the difference between maximal infected rate and bed rate, calculated as the number of beds per 100,000 persons. When the total number of field hospital beds was plotted against (infected rate – bed rate) **(Fig 3B)**, indeed the correlation is stronger than that against any other statistics, followed by state population and accumulative confirmed cases **(Supplemental Fig 3)**. This phenomenon should be largely credited to the IHME projection model, which provided state-specific guidelines in preparing for the upcoming surge.

**Figure 3:**
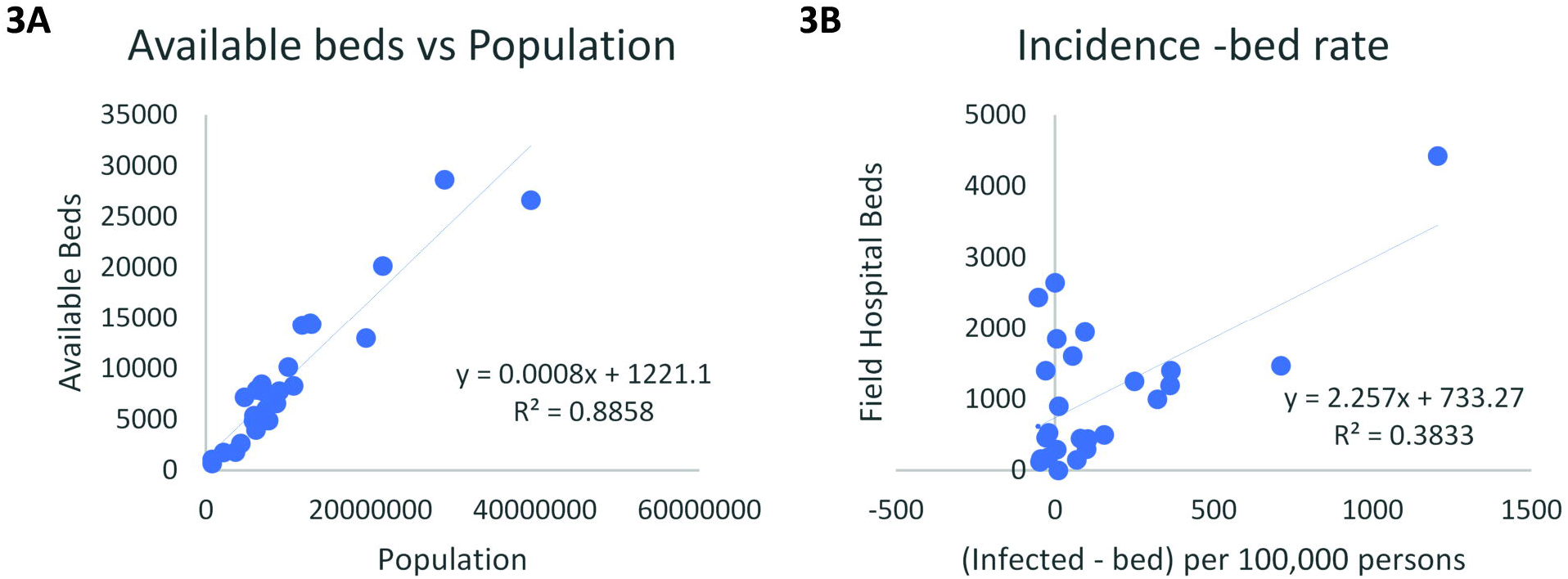
The number of field hospital beds is mostly correlated to incidence minus bed rate, a proxy for the additional beds required predicting using SIR model. **3A:** The number of available beds before COVID-19 is proportional to the population in each state. 3B: The number of field hospital beds in each state is mostly correlated to incidence minus bed rate.

### Different types of field hospitals were utilized differentially

When the field hospitals had been constructed with the right capacity and ready to receive patients around demand peak, the final question came down to: what purpose should they serve? Should they treat routine non-COVID-19 patients—such as those with kidney failure, heart disease, or cancer—to restore normal operation of the healthcare system, or should they take up COVID-19 patients when there is not enough space in the local hospitals? Should there be intensive care units or for mild-symptom and recovering patients only? While the local hospitals have been toiled by the flood of COVID-19 patients, it seemed like a cost-effective plan to designate field hospitals as overflow facilities to care for mild and recovering COVID-19 patients, where neither extra medical equipment, such as ventilators, nor specially trained staff were required. However, the follow-up news coverage may suggest otherwise **(Table 3)**.

**Table 3:** News coverage on opened field hospitals^70-77^. Positive reports are marked in green and negative in red. Checkmarks indicate the commentaries the field hospitals received. Blank indicates no news coverage.

| Hospital Name | COVID | Military | Beds | Opening Date | Utilized well | COVID tested | Low admission | Strict transfer | Closed |
| --- | --- | --- | --- | --- | --- | --- | --- | --- | --- |
| Central Park Field Hospital | Yes | No | 68 | 4/1 | x |  |  |  |  |
| Boston Convention and Exposition Center | Yes | No | 1000 | 4/10 | x |  |  |  |  |
| USNS Mercy | No | Yes | 1000 | 3/27 |  | x | x |  | x |
| USNS Comfort | No | Yes | 1000 | 3/30 |  | x | x |  | x |
| San Diego Convention Center Shelter | No | No | 400 | 4/1 |  | x |  |  |  |
| Javits New York Medical Station | Yes | Yes | 1000 | 4/4 |  |  | x | x |  |
| CenturyLink | No | Yes | 250 | 4/6 |  |  | x |  | x |
| TCF Center Field Hospital | Yes | Yes | 1000 | 4/11 |  |  | x | x |  |

As of end of April, among the 75 surveyed field hospitals, 43 opened, 29 remained closed or under construction, and 3 was canceled; out of the 43 opened field hospitals, 2 had positive news reports, 6 negative, and the rest with no additional follow-up news coverage **(Fig 4A)**. The major concerns included 1) COVID-19 outbreak in a non-COVID-19 facility, 2) low admission rate, and 3) strict transfer rules to accept patients from local hospitals. When the field hospitals were not required any more or could not contain the outbreak, patients would be discharged, and hospital shut down. The relationship between the negative commentary categories is depicted in the Venn diagram **(Fig 4B)**.

**Figure 4:**
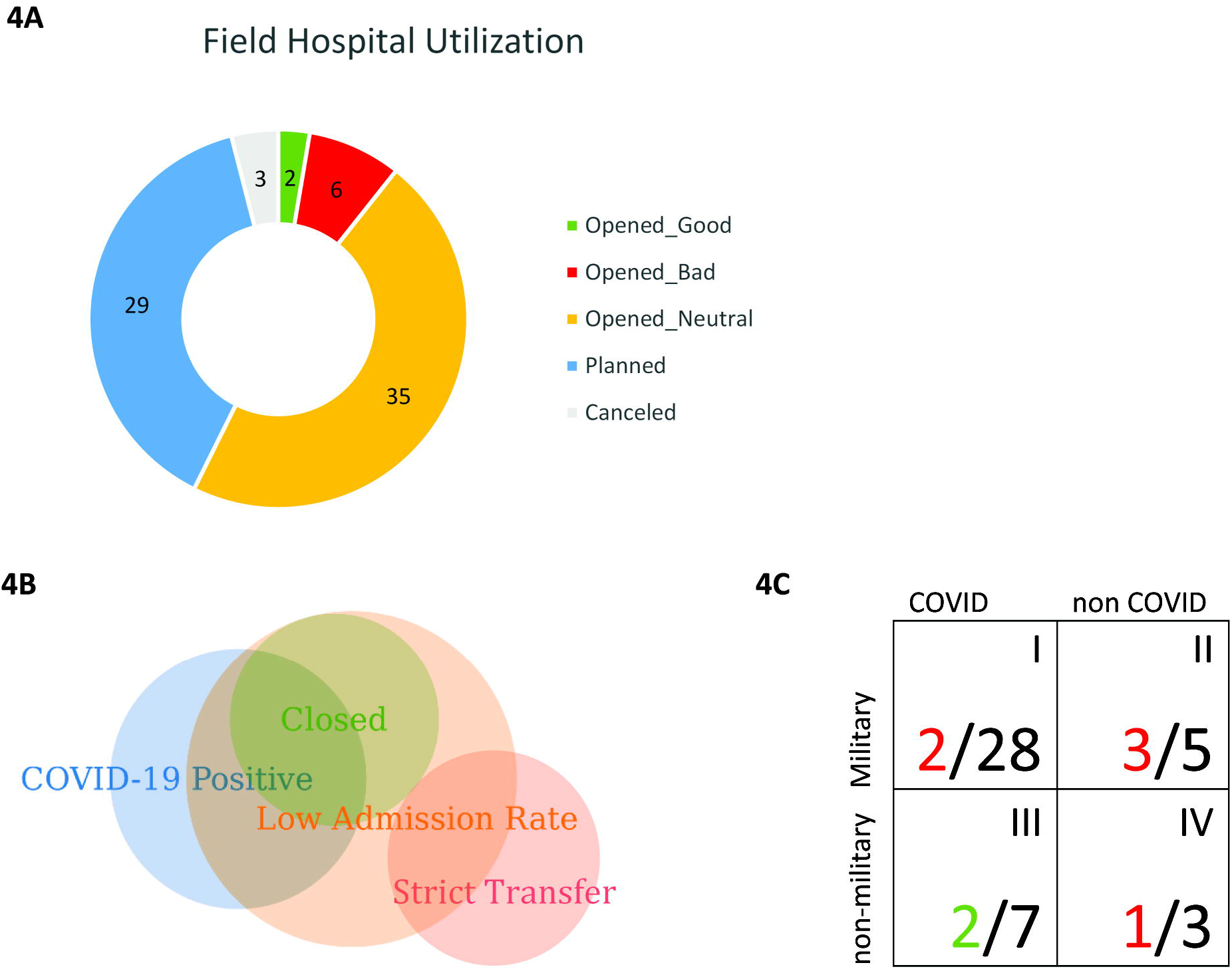
The effectiveness of decision making and the utilization of resources. **4A:** The status of all field hospitals. **4B:** Reasons of negative news report on the opened field hospitals. No conflicting reports were found. **4C:** Positive and negative news coverage assigned to different quadrants of opened field hospitals. Red indicates negative commentaries and green positive. The numerator is the number of reported field hospitals, and denominator the total number of opened hospitals in each category. Each quadrant is labeled with Roman numerals on the upper right corner.

When looking at the **Table 3** more closely, a pattern could be found between the property of field hospitals and the commentary they received. Therefore, we divided up the opened field hospitals into four quadrants depending on the patients they received (COVID-19 vs non-COVID-19), and the affiliation (military vs non-military). Then the reported field hospitals are assigned to each quadrant **(Fig 4C)**. The non-military-built field hospitals are mostly affiliated or sponsored by local healthcare systems and receive direct transfers whereas the larger scaled military-built field hospitals are mostly freestanding facility accepting patients from the entire region. Therefore, it is an indicator of how well the field hospitals are integrated into the local healthcare system.

Javits Medical Center and TCF Center fell into quadrant I (COVID-19, military) and they both had low admission rate due to strict transfer rules. Even though many patients requested transfer, only a small portion was granted admission. The two *USNS* cruises and CenturyLink belong to quadrant II (non-COVID-19, military) and they all had low admission rate. Both *USNS* cruises had staff or patients tested positive for COVID-19. *USNS* Comfort discharged all patients and left New York after 3 weeks of service and *USNS* Mercy dispatched crew members to assist local healthcare facilities. CenturyLink in Seattle received 0 patients 3 days after opening and was soon closed. San Diego Convention Center Shelter in quadrant IV also had 2 confirmed COVID-19 cases among the homeless. The two well utilized field hospitals—Central Park Field Hospital and Boston Convention Center—both fall into quadrant III (COVID-19, non-military). Central Park Field Hospital, especially, is more like an overflow ward of Mt. Sinai Hospital with full capacity to provide intensive care. It admitted 142 patients one week after opening and hospitalized at least 50 patients at all time.

In general, the non-COVID and/or military-build field hospitals appeared less utilized than planned. However, from the perspective of public health, the low utilization rate can have mixed implications.

## Discussion

### Low utilization rate of field hospital could be due to various factors

In response to the COVID-19 pandemic, many countries have built makeshift hospitals. Like many field hospitals in the U.S., the NHS (National Health Service) Nightingale Hospitals in the UK also treated very few patients. The Nightingales are also designed as overflow hospitals when the surge does occur. Therefore, the underutilization demonstrate that the local health systems had coped with the extra pressure COVID-19 brought^14^. Similarly, the low utilization rate in the U.S. field hospitals, which are also built as the last resort during the medical crisis, could be due to that public health interventions, such as social distancing and wearing masks, are effective to “flatten the curve” and the infected population did not grow to the predicted value^15^. According to the IHME model, the estimated maximal daily infections in the U.S. were 260K on March 29^th^, however the actual number peaked on April 8^th^ with 32K confirmed cases^5^. Consequently, the field hospitals prepared for the surge were no longer needed.

On top of the lowered number of infected cases, procedural and socioeconomic factors further decrease the utilization of field hospitals. CDC encourages patients with mild symptoms to practice self-isolation and recover at home, which decreases the proportion of infected cases entering the medical system^16^. In addition, loss of health insurance due to unemployment and confusion of coverage also discourage patients to seek medical attention when symptoms are mild^17-18^. When patients are hospitalized but do not need intensive care, the transfer qualification, specifically in Javits Medical Station and TCP Center Field Hospital, again strictly limits the number of patients admitted to the field hospital even though large volume of patients requested for transfer^75,78^.

### COVID-19 intensive care should be prioritized in the epicenter

The most significant difference between the U.S. and UK field hospitals is that the NHS prioritized the COVID-19 patients needing critical care over mild and recovering patients, with the non-COVID-19 patients coming last. The most important medical center, the ExCel Nightingale, specifically was commissioned with a purpose to take up unconscious, ventilated patients who could not take direct referrals from the community. It was not a “step down” facility, but rather an alternative. Whereas in the U.S., most COVID-19 field hospitals only take mild and recovering patients and had low admission rate. In contrast, the Central Park Field Hospital, equipped with intensive care unit have filled the beds with patients and are running around the clock. Comparatively speaking, it may be a better idea to accentuate the resources on smaller number of intensive care beds instead of on large amount of non-intensive care beds, when most patients can recover without hospitalization^19^.

### Non-intensive COVID-19 field hospitals can serve as entry instead of step-down facilities

The FangCang field hospitals built in Wuhan, China were also designed for mild and recovering COVID-19 patients but had high occupancy. The major difference is that instead of taking patient outflow, FangCang field hospitals triage and quarantine COVID-19 positive patients before the more severe individuals are transferred to designated healthcare facilities^20^. Not only can this workflow free up the space in local hospitals, the confirmed cases can be properly quarantined to prevent further in-person transmission. Whereas many COVID-19 patients with mild symptoms were self-quarantined at home and may not strictly follow the rules, group quarantine patients in field hospital appears a more effective way to utilize the resources.

## Conclusion

When the accumulative confirmed cases surged immensely in mid-March, the affected states have properly responded to the COVID-19 outbreak by promptly building field hospitals to supply additional beds predicted by the IHME model. While many hospitals remained closed to reserve resources, 43 field hospitals opened since late March. The number of follow-up news report on the operation of field hospitals is low, so the conclusion may not be definitive. From what we have gathered, low utilization rate is common. It could be due to a combination of factors, such as non-COVID-19 patients reluctant to seek medical assistance and the outbreak being contained.

By comparing the field hospitals across different categories in the U.S. as well as those in other countries, there could be multiple ways to improve the utilization of medical resources during the pandemic. First, more resources could have been concentrated on intensive care. These field hospitals can be paired with local healthcare systems to allow efficient transfer. Next, non-critical COVID-19 field hospitals can triage confirmed patients and group quarantine them to prevent further transmission. Last but not the least, the non-COVID-19 field hospitals can be kept to minimum and strict screening must be followed to prevent cross-transmission.

## Data Availability

All stated in the context.

## Acknowledgement

The authors would like to thank Ensheng Dong from Center for System Science and Engineering at the Johns Hopkins University for his comments, and the three anonymous reviewers for their insightful suggestions and careful reading of the manuscript. This research was supported by COVID-19 Crisis Relief Program, initiated by the Hopkins Club for Innovation and Entrepreneurship, an alumni and faculty-led 501(c)3 organization in Baltimore, Maryland.

